# Design Of A Rapid And Reversible Fluorescence Assay To Detect Covid-19 And Other Pathogens

**DOI:** 10.1101/2020.10.02.20196113

**Authors:** V. Siddartha Yerramilli, Suzanne Scarlata

## Abstract

We describe a rapid and reusable biophysical method to assay COVID-19 and other pathogens. The method uses fluorescent sensors (i.e. molecular beacons) designed to detect COVID-19 RNA or any RNA of interest, concurrent with an internal control without the need for amplification. The molecular beacons are stem-loop structures in which a ∼10 nucleotide loop region has the complementary sequence of a region of the target RNA, and a fluorophore and quencher are placed on the 5’ and 3’ ends of the stem. The energy of hybridization of the loop with its target is designed to be greater than the hybridization energy of the energy of the stem so that when the beacon encounters its target RNA, the structure opens resulting in dequenching of the fluorophore. Here, we designed a COVID-19 beacon that is completely quenched in its native form and undergoes a 50-fold increase in fluorescence when exposed to nanomolar amounts of synthetic viral oligonucleotide. No changes in intensity are seen when control RNA is added. A control beacon to a human GAPDH RNA, chosen for its high levels in saliva, behaved similar to the COVID-19 beacon. This increase in fluorescence with beacon opening can be completely reversed upon addition of single stranded DNA complementary to COVID-19 beacon loop region. Beacons can be attached to an insert matrix allowing their use in concentrated form and can be made from morphilino oligonucleotides that are resistant to RNases. We present an analysis of the parameters that will allow the development of test strips to detect virus in aerosol, body fluids and community waste.

**Statement of significance:** A platform for reusable and rapid detection of COVID-19 RNA and other pathogenic RNAs without the need for amplification or sophisticated instrumentation in a complex environment is described.

## Introduction

Stopping the spread of COVID-19 has been greatly inhibited by the ability to rapidly and inexpensively assay the presence of the virus in an individual and in a community. While most assays use nasal swabs, more recent assays use saliva which may contain high levels of virus in an infected individual [1]. Assessment of community infection can be determined by continuous monitoring of sewage from dormitories, apartment complexes and other residences, which will alert the need for individual testing (see https://www.bbc.com/news/science-environment-53257101).

Here, we describe a simple, reusable fluorescent assay for COVID-19 RNA. This assay has the potential to simultaneously detect and monitor multiple RNAs in real time in a bar code format. The method uses molecular beacons [2]. Molecular beacons are oligonucleotides arranged in a stem-loop ‘lollipop’ structure. At the bottom of the stem is a fluorophore on one end (either the 5’ or 3’) and a quencher on the other. The nucleotides in the loop are designed to be complementary to a target a specific mRNA so that when the beacon is exposed to its target mRNA, hybridization occurs. Hybridization opens the stem, pulling the quencher away from the fluorophore resulting in a large increase in fluorescence (see **Fig. 1A**).

**Figure 1.**
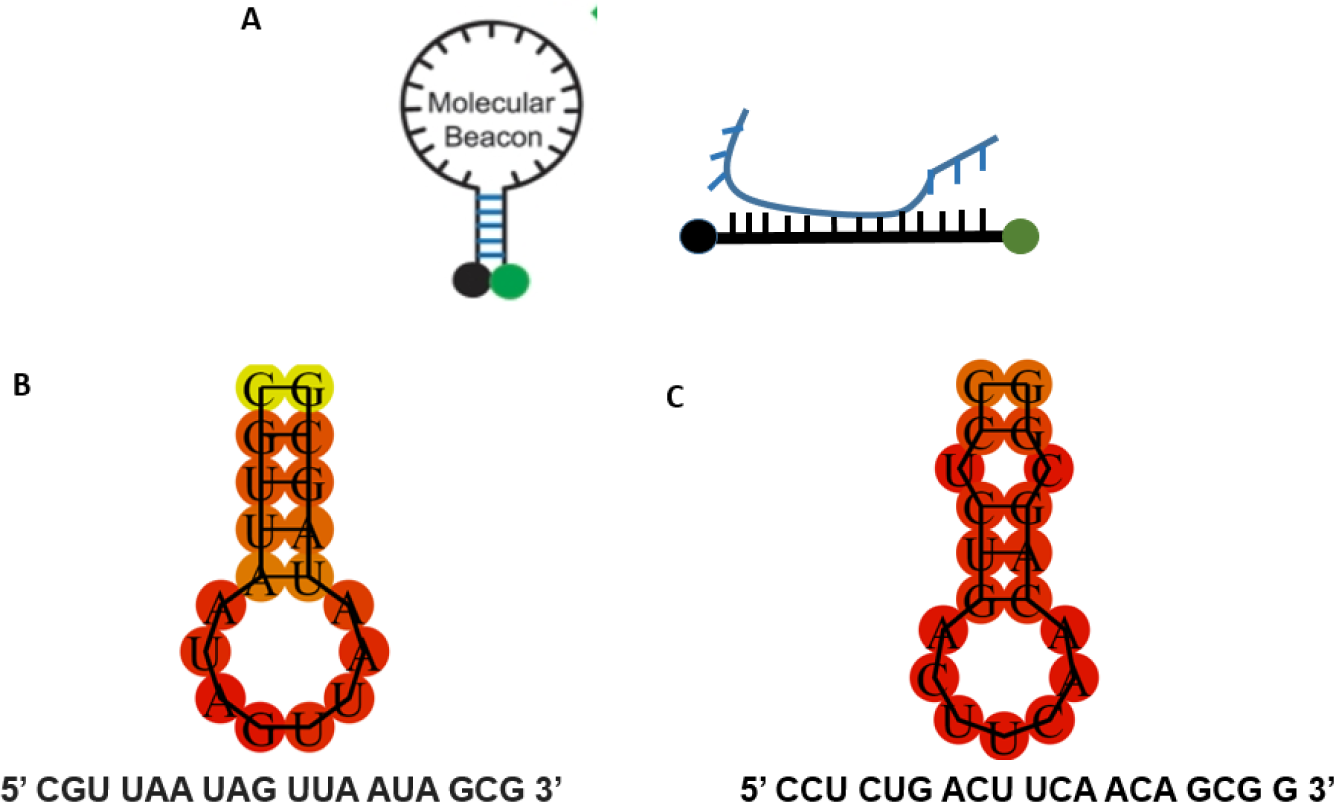
**A.** *Left-* diagram of a molecular beacon showing the stem-loop (closed) structure. The close proximity of the fluorophore (green circle) to the quencher (black circle) eliminates all fluorescence. *Right*-upon encountering target mRNA (blue) the beacon opens and becomes fluorescent. **B**. Beacon used for COVID-19. **C**. Beacon used for GAPDH.

Previously, our lab has used molecular beacons to isolate stem cells induced to express exogenous genes [3]. In those studies, we designed a molcular beacon with a green fluorophore that targeted an exogenous gene that codes a choloride channel that helps transform stem cells into a cardiac pacemaker lineage. The studies necessitated the use of a tracking probe that was incorporated into the beacon between the stem and the loop to identify cells containing the beacon by red fluorescence, and cells containing the mRNA of the gene of interest by green fluorescence. Cells expressing the target gene were then separated by fluorescence activated cell sorting. In recent months, a molecular beacon detection system for COVID-19 has been produced although this system is solution based an requires amplification and specialized reagents. In this study, we show how molecular beacon can be applied in a simple, rapid and reusable platform to simultaneously detect COVID-19 and other pathogens in complex environments without the need for specialized instrumention that might be beneficial for use in underdeveloped nations.

## Methods

### Gene sequences

The sequence for the COVID envelope protein was derived [4] and later verified using BLAST. The sequence for GAPDH was beacon was obtained randomly from the GAPDH mRNA and further verified using BLAST.

### Materials

The beacons labeled with FAM at te 5’end and Iowa-BHQ (black hole quencher) at the 3’end were purchased from IDT Technologies. The single stranded DNA oligonucleotides were obtained from Eton Biosciences. The sequences used for the oligonucleotides Beacons and oligonucleotides were dissolved into stock solutions and further diluted using DEPC water.

### Fluorescence measurements

All measurements were conducted on Cary Eclipse fluorometer (Varian). Fluorescence was measured in a quartz cuvette where the excitation wavelength was 490 nm and emission was 520 nm. The data was exported from the manufacturer’s software into graphing softwares such as Excel (Microsoft) and Prism (GraphPad).

### Saliva collection

Whole-mouth saliva samples were self-collected in DNase- and RNase-free Falcon tubes (50 mL). The volumes collected ranged from 5 ml to 10 ml. If the samples are not processed for RNA extraction immediately, the samples were frozen and stored at −80 °C until further analysis. The individual has been tested negative for COVID-19 using SARS-CoV2 Real-time Reverse Transcriptase (RT)-PCR Diagnostic Assay prior to sample collection.

### RNA extraction

Whole cell RNA was obtained from HeLa cells (ATCC) with RNAeasy extraction kit (Qiagen) based on the manufacturer’s instruction. RNA was isolated from human saliva by boiling human saliva specimen with 1X TE buffer at 95C as described earlier [5].

## Results

### Beacon Design

Ideally, beacons should be designed to detect unique sequences in the 3’ untranslated region of the mRNA. This length is long enough to give stable hybridization with target mRNA and short enough so that stable secondary structures within the loop do not occur, and thus the optimal length of the loop is usually ∼10 nucleotides. Similarly, the optimal length of the stem is ∼5 base paired nucleotides on the 5’ and 3’ ends which can be longer or shorter to enhance stability. This short length stabilizes the beacon structure and can be easily destabilized by hybridiation of the loop region with the target mRNA.

We designed a COVID-19 beacon based on the primers described by Kim et. al [4] that was in turn based on the SARS-CoV genome detailed in Paraskevis et al [6]. The SARS-CoV-2 mRNA encodes a range of structural and non-structural proteins that can be targeted, i.e. spike glycoprotein, envelope protein, membrane glycoprotein, nucleocapsid protein and RNA-dependent RNA polymerase. We designed a beacon corresponding to the mRNA encoding for the envelope protein region of SARS-COV-2 that was verified to not match any in the human genome using BLAST. Based on the selected loop region, the stem nucleotides were chosen to be 5’ CGU UAA UAG UUA AUA GCG using the pairing energies calculated with RNAfold software[7]. The free energy of the thermodynamic ensemble is -1.82 kcal/mol and the melting temperature is 36.7° C. (**Fig 1B**).

As a control, we also designed a beacon to GAPDH whose mRNA expression levels in saliva have been used extensively as a reference gene [8]. The sequence and energetics associated with this beacon are 5’ CCU CUG ACU UCA ACA GCG G whose predicted free energy of the thermodynamic ensemble is -2.66 kcal/mol and the Tm is 55°C. (**Fig**.**1C**). For this proof-of-concept study, we attached FAM and BHQ at the 5’ and 3’ ends of the beacons, and note that responses can be optimized with probes optimized to the specific conditions and instrumentation.

### Beacon performance

We first tested the response of the beacons dissolved in RNAse free water on an analog fluorometer. The COVID-19 beacon had a fluorescence intensity comparable to background. However, upon addition so synthetic COVID-19 mRNA, the fluorescence at 488nm showed a large and reproducible ∼50 fold increase (**Fig. 2A**). The GAPDH beacon had a similar response as the COVID-19 beacon as expected from the similar energetics of the oligonucleotide sequence and the attachment of the same probes (**Fig. 2B**).

**Figure 2.**
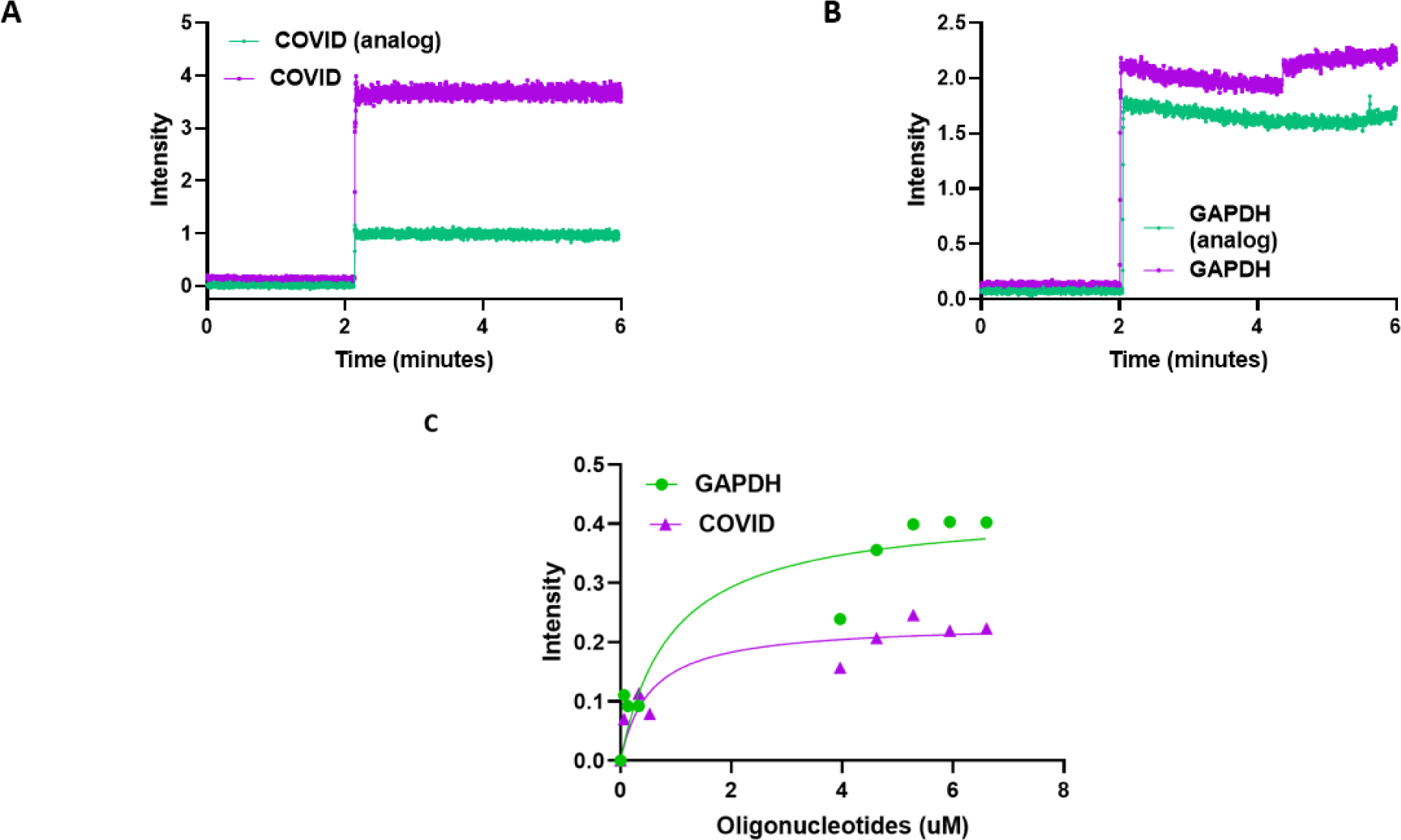
**A.** COVID-19 beacon (final concentration 400 nM) was added to 1 μM of DNA oligos that is analogous to (COVID analog CGTTAATAGTTAATAGCG) or complementary to COVID-19 beacon (COVID GCAATTATCAATTATCGC) in a cuvette in a fluorometer **B**. GADPH beacon (final concentration 400 nM) was added to 1 μM of DNA oligos that is analogous to (GAPDH analog CCTCTGACTTCAACAGCGG) or complementary to GAPDH beacon (GAPDH GGAGACTGAAGTTGTCGCC) in a cuvette in a fluorometer **C**. COVID and GAPDH DNA oligos were titrated into 250 nM of COVID-19 and GAPDH beacons respectively. The fluorescence measurements were then fitted using a curve fitting function to calculate the Kd of the RNA-DNA interaction. All fluorescence measurements were taken at an excitation wavelength of 490 nm and emission wavelength of 520 nm.

Titration of the target oligonucleotide with beacons gave us a K_d_ of 532 nM for COVID and 910 nM for GAPDH when added to 250 nM of beacons giving us a response factor (i.e. the amount of opening of the beacon with RNA) of 1:2 for the COVID-19 beacon and less than 1:4 for the GAPDH beacon (**Fig. 2C**).

Beacon responses were tested in two biological conditions, saliva and HeLa cells. For saliva, samples were heated in TE buffer to expose RNA [5]. We note that RNA can also be released from COVID particles by other methods including heat. When added to the saliva samples, only background fluorescence from the COVID-19 beacon was detected indicating that the closed, non-fluorescence beacon strucutre remains intact in a cellular environme Alternately, the GAPDH beacons showed a high level of fluorescence indicating the presence of endogenous GAPDH RNA. When the samples were spiked with oliognucleotides to COVID-19 the fluorescence increases drastically (**Fig. 3C**).

**Figure 3.**
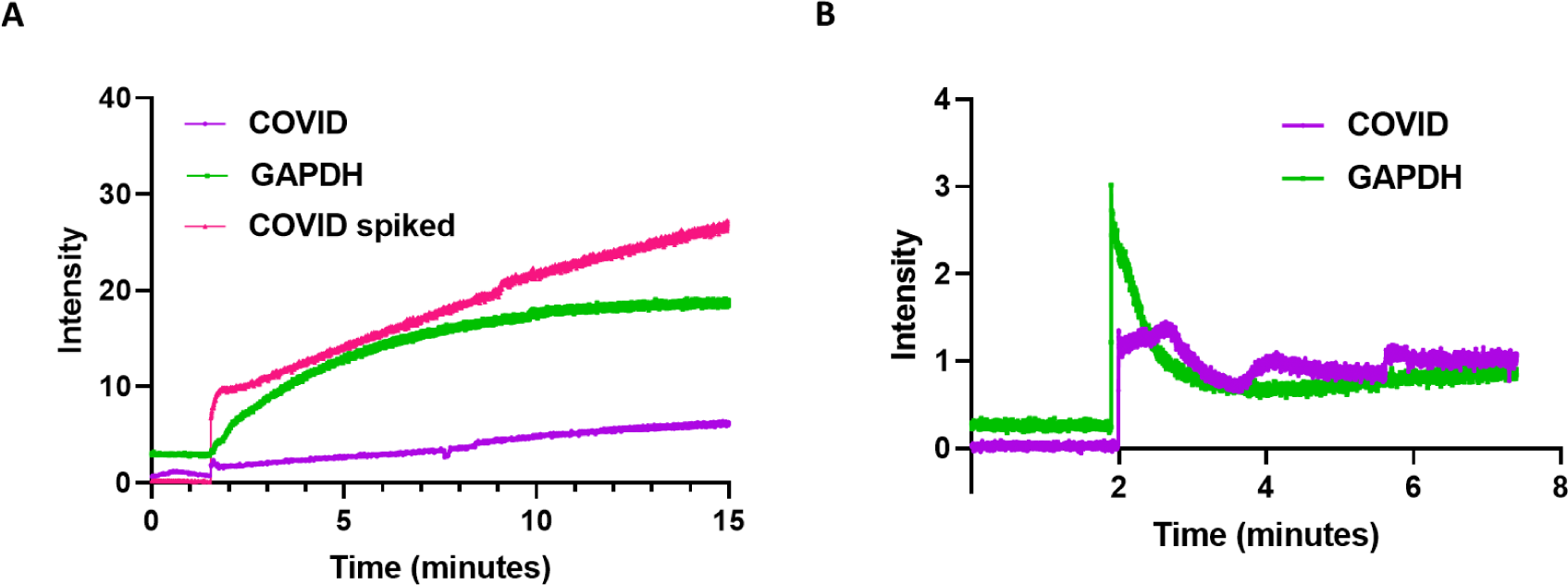
**A.** Human saliva sample boiled in TE buffer was added to 400 nM COVID-19 or GAPDH RNA beacons in a cuvette and then measured in a fluorometer. The saliva sample was then spiked with 20 μM of COVID DNA oligo before being added to 400 nM COVID-19 RNA oligo and measured similarly. **B**. RNA extracted from HeLa cells (190 ng/μl) was added to 400 nM COVID-19 or GAPDH RNA beacons in a cuvette and then measured in a fluorometer. **C**. COVID-19 beacon (final concentration 400 nM) was added to 1 μM of COVID or COVID analog DNA oligos in a cuvette in a fluorometer and measured at temperatures ranging from 20 °C to 60 °C. All fluorescence measurements using an excitation wavelength of 490 nm and emission wavelength of 520 nm.

In another experiment, we added beacons to COVID-19 or GAPDH to RNA extacted from HeLa cells. As expected, no fluorescence was observed when the COVID-19 beacon was added, while a small but measureable fluorescence was seen for the GAPDH beacon due to hybridation with endogenous mRNA(GAPDH). (**Fig. 3B**). Both beacons showed a substantial increase in fluorescence. Interestingly, the fluorescence from the GAPDH beacon showed a spike and rapid decreased that leveled at a high level most likely due to internal mechanisms, the fluorescence from the COVID-19 beacon remained relatively stable possible due to a lack of specific degradation mechanisms.

### Beacon stability

We wished to determine the limitations of the beacons to environmental factors. While the beacons were found to be insensitive to proteases, unfolding of the beacons can be seen at high salt and temperatures. In **Fig. 4** we show the behavior of the beacons alone or with oligonucleotides in solution. Although the calcuated melting temperature is ∼55°C, we find that the midpoint of the melting curves is closer to 48°C. This temperature is unchanged even in the presence of the complementary oligonucleotides since now the hydration energy of this region replaces the loss in energy of the stem.

**Figure 4.**
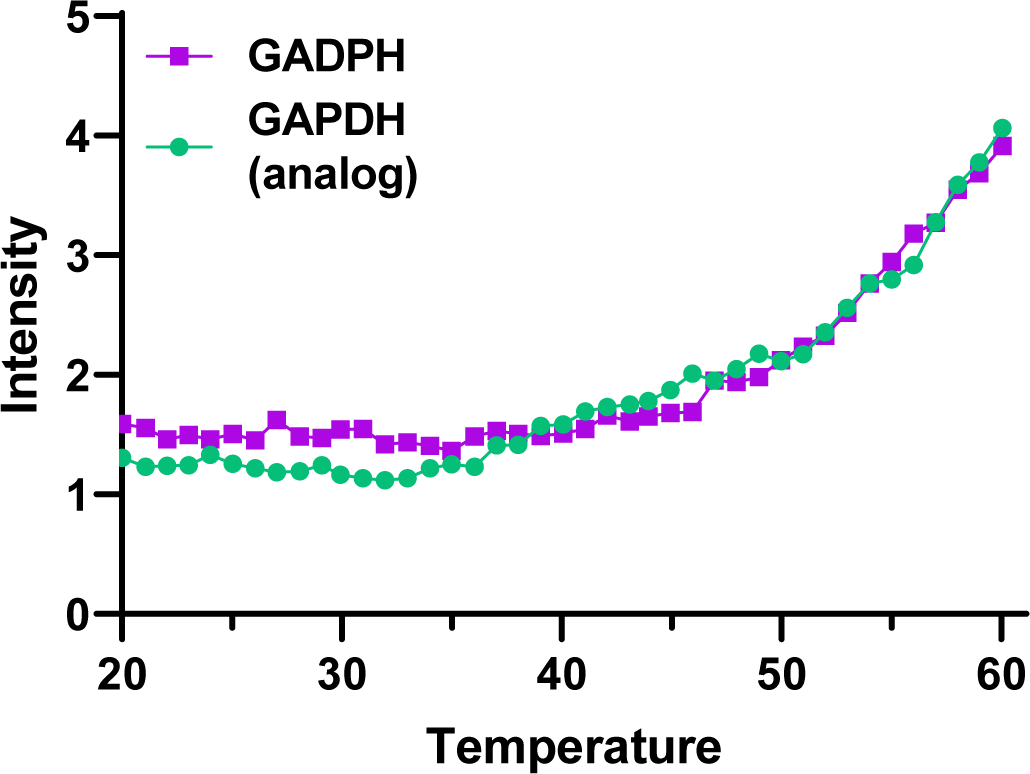
COVID-19 beacon (final concentration 400 nM) was added to 1 μM of COVID or COVID analog DNA oligos in a cuvette in a fluorometer and measured at temperatures ranging from 20 °C to 60 °C. All fluorescence measurements using an excitation wavelength of 490 nm and emission wavelength of 520 nm.

### Viability of using beacons for use in non-clinical settings

The commercial apparatus that uses molecular beacons requires amplification, and specialized components and instrumentation which greatly inhibits their use in underdeveloped nations. Detection of COVID-19 and other pathogens in a non-clinical setting would be possible if beacons were attached to an inert matrix in a concentrated form thereby overcoming the need for amplification, and allow for detection using a simple black light. Beacons can be commercially purchased with a biotin group attached to a thymidine on a stem nucleotide and attached to inert matrices. Thus, the only requirement for an inexpensive assay would be detergent, protease to expose the mRNA from the virus particles, and salt to displace the viral RNA from the beacons and regenerate the matrix or testing strip holding the beacons.

We can easily estimate the optimal density of beacons that should be attached to an insert matrix. Our data show that the response of the beacon is ∼50%. In saliva, the amount of virus has been reported to be several million per milliliter [9] and the amount in aerosol is expected to a few orders of magnitude lower so the needed detection should be ∼500-5000 molecules. Here, we used an FAM probe since fluorescein is inexpensive and has a high quantum yield (0.8) at neutral pH [10]. Noting that the human eye is most sensitive to the fluorescence wavelength of fluorescence and it has been reported that humans can detect as little as one photon [11], detection of 500 photons at ∼500nm by eye should be possible.

To optimize beacon use, it is useful to estimate the density of beacon that must be attached to a test strip. A fully extended beacon would be ∼6 nm and this would be considered the minimal separation to prevent tangling and allow opening and closing without tangling of the molecules. This spacing would also be too far to permit intermolecular quenching or fluorescence transfer. Another potential problem to consider is the small Stokes shift of fluorescein (i.e. 20 nm) that may result in reabsorption of the fluorescence from a neighboring probe [10]. Because the quantum of fluorescein is 0.8, then each reabsorption event will reduce the fluorescence by at least 20%. The distance dependence of trivial reabsorption is R^3^ as compared to R^6^ for Forster resonance energy transfer. Based on our previous characterization of fluorescein [12], then increasing the separation of the beacon molecules by a factor of 10-100 would prevent any untoward optical effects.

By far, the main limitation is the size of the target mRNA [1]. COVID-19 mRNA is large 30kB making its estimated radius of gyration to be close to 1 micron [13], which may cause crowding for beacon separated much more closely. This problem can be resolved by decreasing the beacon density on the assay strip. Alternately posited, by mild digestion of COVID19 mRNA by including small quantities of RNAase in the detergent / protease solution used to release viral particles, and would require the use of beacons be made from morphilino oligonucleotides (see Discussion), or by using a mixture of beacons that detect different regions of a single COVID RNA.

## Discussion

Here, we present guidelines of how a simple and inexpensive COVID-19 assay in complex solutions can be prepared for use in a non-clinical setting. Fluorescence increases from beacon responses signals are rapid and can be reversed by the addition of inexpensive ssDNA with a sequence identical to the loop region, or high salt if attached to a matrix. Beacons can be designed to detect any oligonucleotide of interest. As mentioned, it is best to select of beacon sequences that are unique to the particular RNA, and not found in the human genome, or at least not found in sample materials (see [3]). Our stability studies show that beacons must be kept at temperatures below ∼45°C and other conditions that may promote premature opening. Design of the stem region can be made after calculating the hybridization energy of the loop using freely available software.

We tested the performance of the COVID-19 beacon in solution, saliva and HeLa cells. Instead of using live virus, we spiked solutions with a synthetic COVID-19 oligonucleotide at concentrations below the amount estimated to be in human saliva of an infected individual [14]. In practice, biological samples would have to be mixed with detergent with added salt and possibly protease. Heat treatment has been shown to release viral RNA, and if pure viral RNA is desired when coupling this method to others, treatment with acid can be used [15]. Our data show that the response of the beacon for viral RNA is close to 1:4 and because the beacon can be used in concentrated form (see below) and the viral load of infected patients can range from 10^4^−10^10^ copies/mL [9, 14], we propose that viral RNA can be detected be by eye using a simple black light.

While we focus here on assay development that may benefit underdeveloped nations, beacon technology can be readily adapted for commercial use in individual human diagnostic assays or community detection in sewage, airborne droplets or other fluids. For commercial purposes, substituting natural nucleotides with morphilinos will make beacons resistant to degradation. Morpholinos are identical to regular oligonucleotides with a phosphorodiamidate group instead of phosphate making them resistant to degradation by naturally occurring DNAs and RNAs [16], and morphilino beacons perform similar to DNA/RNA beacons [17]. Importantly, RNA or morphilino beacons can be chemically attached to streptavidin and immobilized on inert matrices using the method described by Bao and corworkers [18]. As discussed, attachment will allow dense concentration of beacons to COVID-19 in a band pattern or barcode at different concentrations, to other pathogenic RNAs to allow for differentiation of different infections, and simultaneous detection of a standard control RNA, such as GAPDH.

## Data Availability

All primary data will be available upon request

## Acknowledgements

This work was supported by NIH GM11687

## Author Contributions

SS initiated the study, designed experiments and wrote the paper. VSY helped designed the experiments, carried out the experiments, analyzed the data and helped write the paper

## Notes

### Competing Interest Statement

The authors have declared no competing interest.

### Clinical Trial

This work was declared exempt from human subject research based on Worcester Polytechnic University's IRB review board.

### Funding Statement

NIH-NIGMS

### Author Declarations

Human Subjects Exemption was issued by the WPI Institutional Review Board, Chaired by Kent Rissmiller, PhD, JD

## References

1. Esbin, M. N., Whitney, O. N., Chong, S., Maurer, A., Darzacq, X. & Tjian, R. (2020) Overcoming the bottleneck to widespread testing: a rapid review of nucleic acid testing approaches for COVID-19 detection, RNA. 26, 771–783.

2. Tyagi, S. & Kramer, F. R. (1996) Molecular Beacons: Probes that Fluoresce upon Hybridization, Nature Biotechnology. 14, 303–308.

3. Guo, Y., Lu, Z., Cohen, I. S. & Scarlata, S. (2015) Development of a Universal RNA Beacon for Exogenous Gene Detection, Stem Cells Translational Medicine. 4, 476–482.

4. Kim, J. M., Chung, Y. S., Jo, H. J., Lee, N. J., Kim, M. S., Woo, S. H., Park, S., Kim, J. W., Kim, H. M. & Han, M. G. (2020) Identification of Coronavirus Isolated from a Patient in Korea with COVID-19, Osong Public Health Res Perspect. 11, 3–7.

5. Ranoa, D. R. E., Holland, R. L., Alnaji, F. G., Green, K. J., Wang, L., Brooke, C. B., Burke, M. D., Fan, T. M. & Hergenrother, P. J. (2020) Saliva-Based Molecular Testing for SARS-CoV-2 that Bypasses RNA Extraction, 2020.06.18.159434.

6. Paraskevis, D., Kostaki, E. G., Magiorkinis, G., Panayiotakopoulos, G., Sourvinos, G. & Tsiodras, S. (2020) Full-genome evolutionary analysis of the novel corona virus (2019-nCoV) rejects the hypothesis of emergence as a result of a recent recombination event, Infect Genet Evol. 79, 104212.

7. Lorenz, R., Bernhart, S. H., Honer Zu Siederdissen, C., Tafer, H., Flamm, C., Stadler, P. F. & Hofacker, I. L. (2011) ViennaRNA Package 2.0, Algorithms Mol Biol. 6, 26.

8. Kumar, S. V., Hurteau, G. J. & Spivack, S. D. (2006) Validity of messenger RNA expression analyses of human saliva, Clin Cancer Res. 12, 5033–9.

9. Yoon, J. G., Yoon, J., Song, J. Y., Yoon, S. Y., Lim, C. S., Seong, H., Noh, J. Y., Cheong, H. J. & Kim, W. J. (2020) Clinical Significance of a High SARS-CoV-2 Viral Load in the Saliva, J Korean Med Sci. 35, e195–e195.

10. Albani, J. R. (2004) Chapter 3 - Fluorophores: Descriptions and Properties in Structure and Dynamics of Macromolecules: Absorption and Fluorescence Studies (Albani, J. R., ed) pp. 99–140, Elsevier Science, Amsterdam.

11. Tinsley, J. N., Molodtsov, M. I., Prevedel, R., Wartmann, D., Espigulé-Pons, J., Lauwers, M. & Vaziri, A. (2016) Direct detection of a single photon by humans, Nature Communications. 7, 12172.

12. Runnels, L. W. & Scarlata, S. F. (1995) Theory and application of fluorescence homotransfer to melittin oligomerization, Biophysical Journal. 69, 1569–83.

13. Gopal, A., Zhou, Z. H., Knobler, C. M. & Gelbart, W. M. (2012) Visualizing large RNA molecules in solution, RNA. 18, 284–299.

14. Wyllie, A. L., Fournier, J., Casanovas-Massana, A., Campbell, M., Tokuyama, M., Vijayakumar, P., Geng, B., Muenker, M. C., Moore, A. J., Vogels, C. B. F., Petrone, M. E., Ott, I. M., Lu, P., Lu-Culligan, A., Klein, J., Venkataraman, A., Earnest, R., Simonov, M., Datta, R., Handoko, R., Naushad, N., Sewanan, L. R., Valdez, J., White, E. B., Lapidus, S., Kalinich, C. C., Jiang, X., Kim, D. J., Kudo, E., Linehan, M., Mao, T., Moriyama, M., Oh, J. E., Park, A., Silva, J., Song, E., Takahashi, T., Taura, M., Weizman, O.-E., Wong, P., Yang, Y., Bermejo, S., Odio, C., Omer, S. B., Dela Cruz, C. S., Farhadian, S., Martinello, R. A., Iwasaki, A., Grubaugh, N. D. & Ko, A. I. (2020) Saliva is more sensitive for SARS-CoV-2 detection in COVID-19 patients than nasopharyngeal swabs, medRxiv, 2020.04.16.20067835.

15. Wozniak, A., Cerda, A., Ibarra-Henriquez, C., Sebastian, V., Armijo, G., Lamig, L., Miranda, C., Lagos, M., Solari, S., Guzmán, A. M., Quiroga, T., Hitschfeld, S., Riveras, E., Ferres, M., Gutiérrez, R. A. & García, P. (2020) A simple RNA preparation method for SARS-CoV-2 detection by RT-qPCR, bioRxiv, 2020.05.07.083048.

16. Summerton, J. & Weller, D. (1997) Morpholino Antisense Oligomers: Design, Preparation, and Properties, Antisense and Nucleic Acid Drug Development. 7, 187–195.

17. Chen, J., Wu, J. & Hong, Y. (2016) The morpholino molecular beacon for specific RNA visualization in vivo, Chemical Communications. 52, 3191–3194.

18. Nitin, N., Santangelo, P. J., Kim, G., Nie, S. & Bao, G. (2004) Peptide-linked molecular beacons for efficient delivery and rapid mRNA detection in living cells, Nucleic Acids Research. 32, e58–e58.

